# Effectiveness of the U-Niko Intervention: Protocol for a cluster randomized controlled trial of a municipal-based tobacco and nicotine cessation intervention for adolescents and young adults

**DOI:** 10.1101/2025.04.11.25325652

**Authors:** Sofie K. Bergman Rasmussen, Christina Bjørk Petersen, M Mette Rasmussen, Nina K Kamstrup-Larsen, Charlotta Pisinger

## Abstract

**Background:** The use of nicotine products among youth is rising, while existing cessation services remain underutilized. Thus, the U-Niko intervention was developed to provide an evidence-based, youth-oriented approach to effective nicotine cessation.

**Methods:** A two-arm cluster randomized controlled trial will be conducted among youth aged 16-25 in 55 municipalities in Denmark. Using the online randomization program Sealed Envelope, we conducted a blinded stratified randomization of 55 Danish municipalities resulting in 27 municipalities in the U-Niko intervention group and 28 municipalities in the control group (tobacco and nicotine cessation recruitment and counseling as usual). The primary outcomes measure the effectiveness of all three focus areas in the intervention group compared to the control group: A) the municipal counselors’ self-efficacy in youth cessation counseling, B) the number of recruited youths for cessation counseling, and C) the self-reported 14-day point prevalence abstinence of youth at six months follow-up. Secondary outcomes are the number of recruited youths in a municipality compared to the previous year, continuous abstinence, and validated 14-day point prevalence abstinence at six months follow-up.

**Discussion:** By evaluating all three focus areas of the U-Niko intervention, this study aims to provide robust evidence for improving youth cessation interventions at a local and national level.

**Trial registration:** The study was registered in ANZCTR (ACTRN12624001470583) on 18/12/2024. The Universal Trial Number is U1111-13-14-6117.

## Introduction

In recent years, youth nicotine use has risen significantly, with nearly 36% of Danish youth (15–29 years) using at least one tobacco or nicotine product (1). While more than half of the young people who smoke and more than 70% of those who use oral nicotine products wish to quit (1), many struggle with high levels of nicotine dependence, making unassisted quitting difficult (2).

The detrimental effects of smoking extend beyond physical health, adversely affecting mental health and contributing to increased school absence in youth (3–5). Further, early exposure to nicotine in any form may yield long-term negative consequences for brain development (6–8) and has been found to function as a “gateway drug” to other psychoactive substances, including cocaine and cannabis (9–11).

Although Denmark offers cost-free, high-quality smoking cessation services, young people underutilize these services and have lower quit rates than adults (12). Group-based smoking cessation counseling is the most frequently used and it is very effective (13); however, municipal counselors report limited experiences with youth-oriented quit services (12) and call for evidence-based strategies to better support young users.

Despite the growing need, there is a lack of high-quality studies on youth tobacco- and nicotine cessation interventions and existing literature is inconclusive (14). However, interventions based on social cognitive theory (15), and group-based smoking cessation counseling (16), seem to be effective in promoting longer-term abstinence in young adults. Regarding cessation of nicotine products (not cigarettes): Low-grade evidence suggests that multi-component interventions that include counseling and peer elements and a text-based cessation program, based on behavioral support, may be effective in the cessation of nicotine products in youth. Pharmacotherapy seems to be well tolerated by young people, but no significant effect has been shown (17,18).

Given the rising use of nicotine products among youth, their difficulties in quitting unassisted, and the underutilization of existing cessation services, there is a pressing need for an effective, evidence-based intervention tailored to young users.

This paper describes the study protocol for a cluster randomized trial evaluating the effectiveness of the U-Niko intervention (’Unge uden nikotin’ – Youth without nicotine), a tailored tobacco and nicotine cessation intervention for youth. Developed for Danish municipalities, which are key stakeholders responsible for cessation efforts, the intervention aims to provide evidence for improving youth tobacco- and nicotine cessation strategies.

## Methods

### Study design

This study is a cluster randomized controlled trial (RCT) including 55 Danish municipalities and aiming at youth aged 16-25. The intervention period will run from January 1^st^, 2025, to December 31^st^, 2025. The municipalities have been randomly allocated to either the U-Niko intervention group or the control group. The youth participants will receive either the U-Niko intervention or a control condition (municipal cessation activities as usual, described under the section *Intervention*) based on the municipality in which they live, go to school, work, or in some other way have their daily activities.

The study is a field trial where all intervention activities are performed by the municipalities, not the researchers. The intervention municipalities do not receive any economic or manpower support from the research group.

Recruitment and cessation counseling are conducted via the municipalities and will occur during the whole intervention year from January 1^st^, 2025, to December 31^st^, 2025. Enrollment and follow-up assessments are conducted via the municipalities in cooperation with the national database on tobacco and nicotine cessation (STOPbasen) and the research group. STOPbasen is a Danish database where most cessation activities in Denmark routinely are registered and have high validity. Data collection is estimated to be complete in June 2026 with results expected to be ready to publish approximately 6-12 months later (Figure 1).

**Figure 1.**
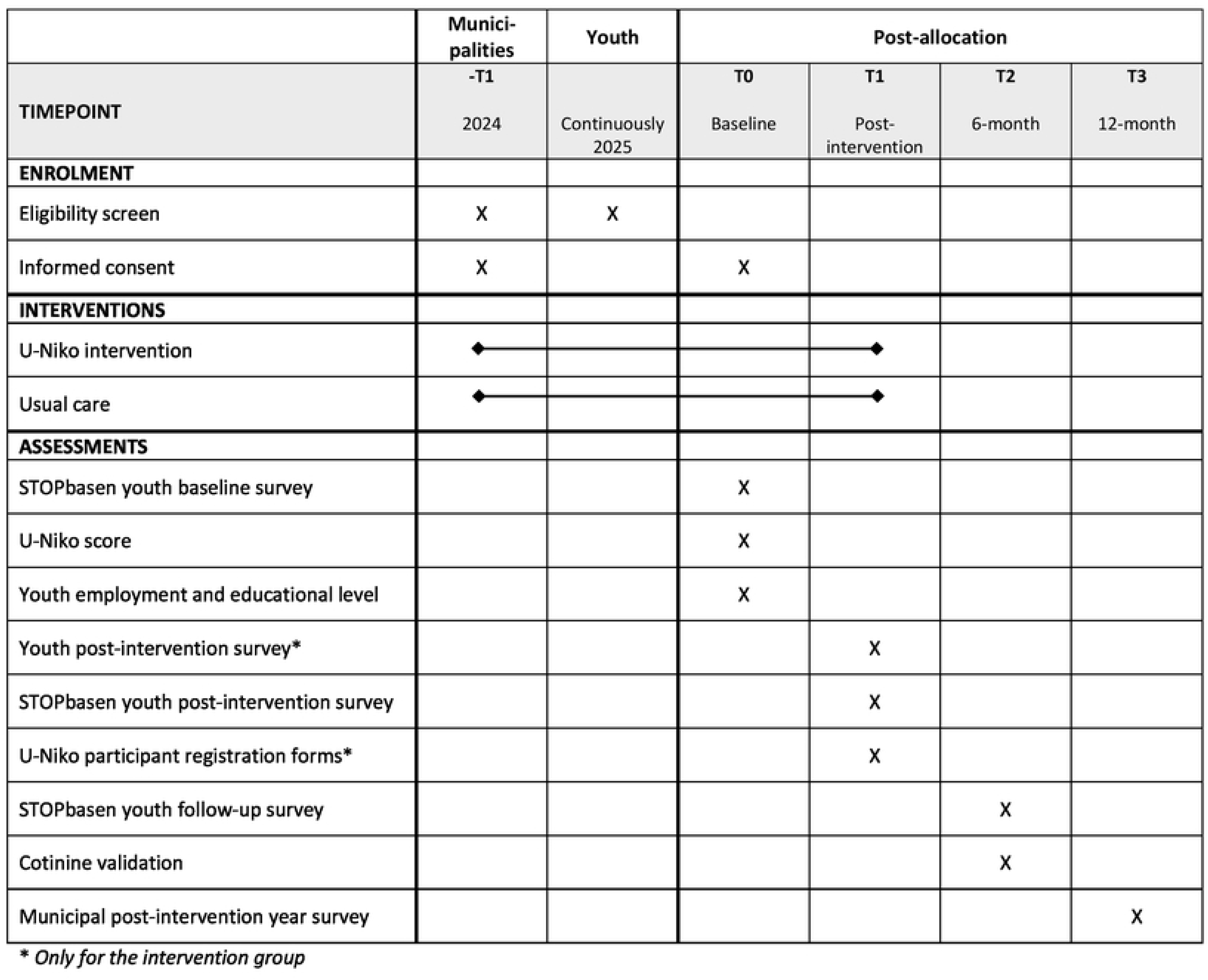
Time schedule of enrolment, interventions, and assessments on participant outcome inspired by the SPRIT 2013 reporting guidelines.

The study is conducted by the Center for Clinical Research and Prevention, Frederiksberg University Hospital, funded by TrygFonden (a non-profit organization), and Health Insurance “danmark” (a non-profit foundation). The study is approved by the Danish National Scientific Ethics Committee (F-24044815), following the General Data Protection Regulation (GDPR) (EU) 2016/679 and was registered in ANZCTR on December 18^th^, 2024 (ACTRN12624001470583). The Universal Trial Number (UTN) is U1111-1314-6117.

All youth receiving support to quit tobacco and/or nicotine products in the intervention and control municipalities will complete a short baseline survey at their first contact with the cessation activities, which will entail a written consent form for the study. The Danish Data Protection Authority and Danish Patient Safety Authority do not require parental permission for minors aged 15 or older.

#### Inclusion criteria

*Eligible municipalities* for the U-Niko intervention are all Danish municipalities willing to work with youth tobacco- and nicotine cessation. Municipalities participating in the feasibility study (n=3) were excluded from the invitation to participate in the effect evaluation.

*Eligible youth participants* for the tobacco and nicotine cessation courses are youth 16 to 25 years old who are currently (daily or occasionally) using at least one tobacco or nicotine product and wish for assistance to quit. Participants will be *excluded* from the study if they do not meet the listed inclusion criteria or fail to provide contact information and informed consent during the enrolment and baseline assessment.

#### Randomization

All municipalities in Denmark that did not participate in the feasibility study (N=95) were invited to register for the trial in March 2024. Forty-three municipalities declined participation and 55 municipalities (58%) of the eligible municipalities accepted, were enrolled and randomized into an intervention group or a control group using the online randomization program Sealed Envelope (Figure 2). During the randomization, municipalities were stratified according to size and inter-municipal cooperation (health clusters), to minimize spill-over effect from the intervention group to the control group. The randomization was blinded. The main reasons for declining participation were that the municipality did not have enough resources to work with young people or that they were already involved in another smoking and nicotine cessation intervention. At the time of randomization, there was great variation in the practices and activities.

**Figure 2.**
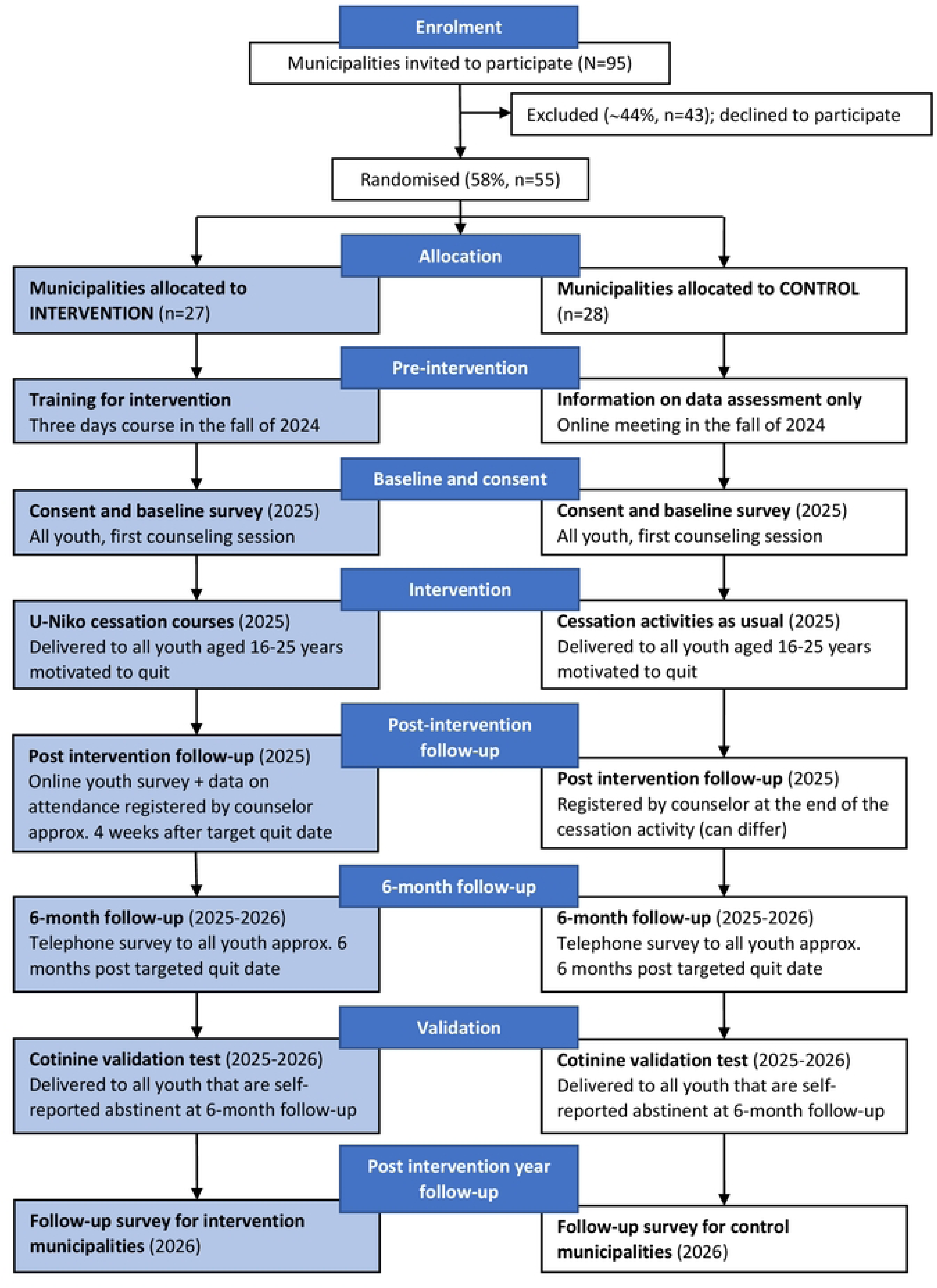
Overview of the intervention enrolment, preparation, trial activities, and data assessment.

### Sample size calculations

The sample size calculations are based on assumptions of difference in self-reported point prevalence abstinence six months after the targeted quit date in the intervention- and control group. Based on data from STOPbasen and international literature (14,19), estimated abstinence under the intention to treat analysis (ITT) is 23% in the control municipalities and 34% in the intervention municipalities six months post-target quit date. Sample size calculations where a municipality is considered a cluster with an inter-correlation coefficient (ICC) of 0.0005 show, that each municipality (both intervention and controls) must recruit at least 11 youth in their cessation activities during the whole intervention year, a total of 605 youth participants.

### Intervention

#### Theory of change

The intervention is tailored to the unique needs of youth users of tobacco and nicotine products. It has been developed based on state-of-the-art research, insights from experienced Danish cessation counselors, and preferences of young tobacco and nicotine users in partnership with Rygestopkonsulenterne ApS (The smoking cessation consultants), a consulting firm with more than 20 years of experience with tobacco- and nicotine cessation counseling (See Appendix for details on the development of the intervention).

The intervention targets three focus areas:

A) **Training** of counselors who assist young smokers and nicotine product users to quit
B) **Recruitment** strategy for youth to tobacco and nicotine cessation services
C) **Cessation** course for youth who use tobacco and nicotine products.

It is hypothesized that A) counselors trained through the U-Niko will report higher skills and confidence in assisting youth quit attempts than those in control municipalities, B) that intervention municipalities will recruit more youth into cessation services than controls, and C) that youth participating in the intervention municipalities’ cessation services will have higher abstinence rates after six months than those in control municipalities (Figure 3). Further, it is expected that there will be a synergistic effect by optimizing all three areas at the same time.

**Figure 3.**
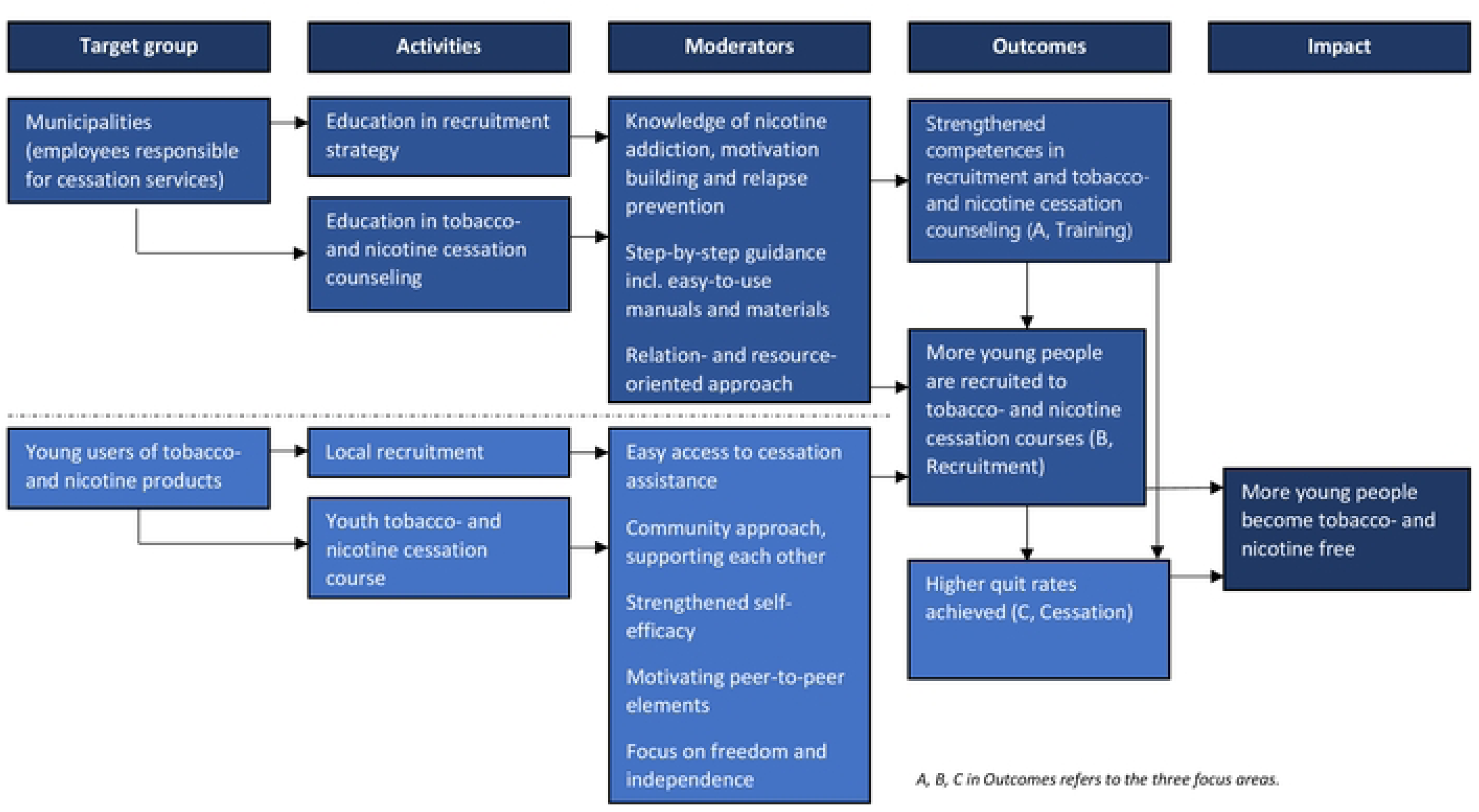
The program theory of change in the U-Niko intervention.

#### The intervention municipalities

The municipalities will receive the U-Niko intervention, including online and physical materials, for all three focus areas. An overview of the key elements of the intervention is provided in Table 1.

**Table 1.** Overview of the U-Niko intervention content.

| Component | Key elements |
| --- | --- |
| A. Training | • Knowledge of nicotine addiction and relapse prevention<br>• Knowledge and concrete tools to work with youth via behavioral and pedagogical approaches<br>• Knowledge of motivation and ambivalence<br>• Basic knowledge of novel tobacco and nicotine products and their harmful effects<br>• Knowledge of the use of the U-Niko score and how to assess physical dependence<br>• Instructing stress reduction exercises |
| **B. Recruitment** | • Step-by-step guidance and tools on how to contact and establish collaborations with youth arenas<br>• Guidance and examples of how to promote the course on tobacco- and nicotine cessation in a motivating way using face-to-face interactions in the youth arena and tools such as videos and posters<br>• Recommendations and examples on how to keep in touch with young people and strengthen their motivation from registration to attendance at the course<br>• Tools and guidance on how to strengthen existing collaboration with a youth arena and how to expand to other arenas year by year |
| **C. Cessation** | • Group-based approach, strong focus on community<br>• Commitment to complete cessation from all tobacco and nicotine products<br>• Behavioral counseling<br>• Strengthening of self-efficacy<br>• Focus on a trust-based relationship, to make all participants feel safe<br>• Peer elements and focus on sharing experiences<br>• Understanding of own addiction and tools on how to overcome cravings<br>• Stress reduction exercises<br>• Text messages to encourage and remind the participants about cessation activities |

*A, B, C in refers to the three focus areas*

##### A. Training

Training in the recruitment strategy is delivered through a 1-day course. Guidance is offered on how to engage with youth (a face-to-face interaction with them in classes or at their workplace, sports club, campus, etc.) and how to motivate them to participate in the course. Videos and posters featuring young peers (role models) sharing their reflections on smoking/nicotine use and their desire to quit are provided.

The youth counselor training is delivered through a 2-day course. The youth counselor training program is aimed at both experienced smoking cessation counselors and individuals with a pedagogical background who have never worked with smoking cessation. It focuses on relational and resource-oriented behavioral techniques, knowledge on nicotine addiction as well as on how to motivate youth to quit tobacco and nicotine and how to prevent relapse. The program focuses on a nonjudgmental and supportive approach. Additionally, the training gives a foundational understanding of novel tobacco and nicotine products and their harmful effects.

##### B. Recruitment

The recruitment strategy is developed as a step-by-step guidance on where to recruit and how to contact youth arenas for recruitment (i.e., schools, workplaces, fitness centers, sports clubs, campuses, barracks, and meeting places for youth) as well as how to encourage them to cooperate. The intervention group will receive free recruitment materials (a video and letters) and guidance on how to recruit in different youth arenas over time.

##### C. Cessation

The course consists of seven meetings planned over eight weeks. The sessions last between 1.5 hours and 45 minutes. Approximately twelve young people are recruited to a group.

The course is anchored around key constructs from social cognitive theory and posits that learning occurs in a social context with a dynamic and reciprocal interaction of the person, environment, and behavior (20). Peer elements are used both in the recruitment and in group counseling. The approach to young people stems from theories from cognitive behavioral therapy (21), as well as narrative engagement theory (22).

The relationship-oriented work is also based on the International Child Development Programme (ICDP) and its eight interaction themes as a tool for a relationship and resource-oriented practice (23). The exercises used stem from mindfulness-based cognitive therapy (24) and mindfulness-based stress reduction (25).

As part of the cessation course, a newly developed score (The U-Niko score, validated by 25 young people, See Appendix 2 for details on the development) is used to measure the physical nicotine dependence. The score has been developed because the nationally used Fagerström score is not suitable for youth since many young people use more than one tobacco and nicotine product, the consumption of e-cigarettes is difficult to measure, and the nicotine content in e-cigarettes and nicotine pouches can vary a lot. The U-Niko score consists of five short questions ranging from 0-9, where 9 being the highest. A score of ≥ 6 indicates high physical nicotine dependence, suggesting that nicotine patches may be beneficial as a part of the cessation process. For individuals aged 18+ with a nicotine patch allergy, cytisine is recommended for those with a score of ≥6.

#### The control municipalities

The municipalities in the control group will continue their recruitment practices and their youth cessation activities “as usual”. The control municipalities can offer individual or group-based counseling, of any duration.

The municipalities are not provided with any information about the intervention or any assistance to improve their recruitment strategies, training of counselors, or youth-oriented cessation activities.

However, the control municipalities will be instructed on how to register baseline and follow-up information and will be offered information on how to use the U-Niko score, as the goal is to compare the young participants’ nicotine dependence uniformly, in the intervention and control groups. Information on how to use the U-Niko score was delivered via an online meeting in December 2024 (Figure 2).

### Evaluation design

#### Measures and data assessment

The data assessment includes data on the effectiveness of all three focus areas of the U-Niko intervention and involves seven different data sources (Figure 1).

##### A. Training

At the beginning of 2026, the year after the intervention year, all municipal counselors from both intervention and control municipalities will receive an online survey examining their self-perceived competencies in working with youth tobacco- and nicotine cessation. Further, the counselors from the intervention municipalities will be asked about their experience with the U-Niko intervention.

**The primary outcome** is *the effect of the training of new youth counselors and* will be measured by assessing the counselor’s self-efficacy in recruiting young people and assisting them to quit and their self-reported perception of and content with their work as youth cessation counselor.

##### B. Recruitment

Data on the number of recruited youths are registered in STOPbasen. Additionally, intervention municipalities are asked to complete a “registration form” for all young people starting in a cessation course. This registration form consists of information on each recruited youth’s participation rate throughout the course.

**The primary outcome** is t*he effect of the recruitment strategy* and will be assessed by comparing the number of recruited youth participants in the intervention municipalities compared with the control municipalities.

**The secondary outcomes** are the *change in the number of recruited youths* to cessation activities between the intervention year 2025 and the previous year 2024, and the *number of recruited youth who participated in the* course in the intervention municipalities.

##### C. Cessation

All participants in both the intervention and control municipalities will, at the first cessation session, complete a baseline questionnaire on contact information, personal characteristics, information on tobacco and/or nicotine use, as well as the U-Niko score. The baseline questionnaire also contains a consent form for participating in the study. The baseline questionnaire will be registered in STOPbasen.

Further, the young participants in the intervention group will be invited to answer a short online survey on their experiences with the intervention after the course has ended (4 weeks post target quit date) regardless of their tobacco- and nicotine status.

Six months after the targeted quit date, all participants (in both the intervention and control group) will be contacted by phone to examine the long-term effect of the cessation offer. A research assistant will contact the participants multiple times by phone as well as send text messages, to decrease the number of missing values at follow-up.

All participants will be informed that for each completed questionnaire (baseline survey, post-intervention survey (only intervention group), and six-month follow-up survey), they will be entered into a raffle with the chance to win a gift certificate worth 1000 DKK (∼134 euros/139 USD). In total, six participants from the intervention municipalities and four from the control municipalities will be drawn to win.

Participants who report being tobacco and nicotine-free at the six-month follow-up will be invited to do a saliva cotinine test to validate their abstinence. The saliva test will be mailed to each participant who has agreed and will be done via an online interview using Teams. The participant will take the test and show the answer to the research staff during the online meeting, discard the test, and receive a gift certificate for 300 DKK (∼40 euros/42 USD), regardless of the outcome of the validation test.

**The primary outcome** is *the effect of the cessation course* will be measured using self-reported point prevalence abstinence rates (not using any tobacco or nicotine product in the last 14 days) six months after the targeted quit date in the intervention municipalities compared with the control municipalities. To assess this, participants will be asked, “Have you used any tobacco or nicotine products in the last 14 days?” (Yes/No).

**The secondary outcomes** are *self-reported abstinence post intervention* (i.e. at the end of the cessation course/counseling), *continuous self-reported abstinence* (from targeted quit date to six-month follow-up), and v*alidated point prevalence abstinence* which will be assessed in all participants who report being tobacco and nicotine abstinent at the six-month follow-up and agree to participate in the cotinine validation process.

#### Additional measures

Additional measures on the participants will be assessed in the baseline survey, including demographic information such as age, sex, gender, socio-economy, the U-Niko dependence score, and product use (type, frequency, duration). Furthermore, data on the employment of the youth participants at baseline will be obtained from the Danish Education Register.

Additional measures for youth cessation counselors will be assessed during the post-intervention municipal follow-up survey. These measures include education and previous experience with tobacco and nicotine cessation activities.

### Data Analysis Plan

Analyses of the effectiveness of the U-Niko intervention will be done by assessing the effectiveness of each of the three focus areas and comparing the control and intervention municipalities. All analyses will be conducted following the intention-to-treat (ITT) principle, where participants will be analyzed in their originally assigned groups regardless of adherence to the intervention. Data will be analyzed using descriptive statistics, bivariate analyses, and multivariate regression models to assess the effectiveness of the U-Niko intervention. Between-group differences (intervention vs. control) and between-individual differences will be assessed using appropriate statistical models.

As data from STOPbasen has shown a high degree of completeness, it is intended to employ a complete case analysis, excluding patients with any missing values in the included variables from the analysis (listwise deletion). Consequently, sensitivity analysis using an as-observed approach will be conducted, including only participants with a valid follow-up. The sensitivity analyses will be performed using the final adjusted model from the main analyses. This approach builds on the assumption that data is Missing Completely At Random (MCAR). However, when it comes to smoking status, data are more likely to be Missing Not At Random (MNAR). Therefore, if warranted, considering the number of missing values and loss to follow-up, a sensitivity analysis using a chained multiple imputation model will be conducted based on the final adjusted model in the main analyses.

## Discussion

This paper describes the study design for evaluating the effectiveness of the U-Niko intervention, a national municipal-based tobacco and nicotine cessation intervention for Danish youth aged 16-25. By integrating insights from field experts, experiences, and preferences of young tobacco and nicotine users, and state-of-the-art research, this intervention aims to serve as an effective supplement to existing cessation services in Denmark.

The study is a nationwide field trial, extending across the country counting 55 municipalities from both rural and urban areas of Denmark. This is a great strength since the intervention will be tested among youth with very different socioeconomic and cultural backgrounds. If the intervention were only tested in certain areas, overall knowledge of its effectiveness for all types of young people would not be attainable.

Another strength is that it is a field trial, not driven by researchers. If the intervention activities prove to be effective, and the municipalities find them helpful, meaningful, and easy to use, they will continue to use them. Thus, there is a high probability of sustainability after the intervention year has ended. By testing the effectiveness of the intervention in 27 different municipal settings, we can acknowledge context-dependent variables such as social, organizational, political, and economic features that influence the intervention effectiveness (26). As more than half of the municipalities in Denmark test the U-Niko intervention, it has great attention, both from the not-included municipalities and from the Health Authorities.

Since the study is a field trial and the intervention occurs in a real-life setting, there are also some uncertainties regarding comparison groups, data registration, and follow-up (27). The youth-oriented cessation activities in the municipalities have been very sparse in the last years, and there is a risk that only a few young persons will be included in 2025, resulting in too low power to detect significant differences between the intervention and the control municipalities. However, in early 2024, the Danish Health Authorities launched a call where municipalities interested in improving their work with youth tobacco and nicotine cessation activities could apply for funding. In total, 58 million DKK (∼7,7 million euros) were allocated to this work and funding must be used between fall 2024 and 2027. This may have a negative impact on the study, as some municipalities will have a much higher activity level than anticipated.

Nonetheless, both intervention (∼44%, n=12/27) and control municipalities (∼54%, n=15/28) have received funding and therefore, the hopefully positive effect of the increased resources will be present in both the intervention and control groups. Therefore it is expected that the calculated sample size requirements on 11 participants during the whole intervention years in each municipality can be accommodated. The analyses of the effectiveness of the U-Niko intervention (recruitment and cessation) will consider that some of the municipalities (more control municipalities) have received extra funding and therefore have more resources.

Another uncertainty is that the municipalities have the responsibility to ensure, that each young participant completes the baseline survey and consent form, which can be forgotten, and follow-up surveys may not be prioritized by the young participants. Further, data rely mostly on self-reported measures which may be subject to reporting biases. However, to motivate higher participation rates in the follow-up activities, all youth participants will enter a competition to win a gift certificate for 1000 DKK (∼134 euros/139 USD) each time they finish a survey. In total, ten youth (six from the intervention group and four from the control group) will win a gift card. Financial incentives to improve youth survey participation rates are known to be very effective (28), but should be used in a way that will not influence the young people’s answers (i.e. only be able to receive the gift certificate if you are tobacco and nicotine-free). Since participating in the competition to win a gift certificate is not dependent on whether the young person still is using tobacco or nicotine, it is anticipated, that it will not result in any biases. Further, all youth who state to be tobacco and nicotine abstinent and agree to participate in the cotinine validation after the six-month follow-up will receive a gift certificate for 300 DKK (∼40 euros/42 USD) regardless of the test outcome. By offering these incentives to participate in the surveys, it is expected that validity will increase, and fewer young participants will be lost to follow-up.

The U-Niko intervention is rather resource-intensive, especially face-to-face counseling. Previous studies have shown that low-intensity interventions might be effective. For example, a text-message-based cessation intervention targeting youth users of e-cigarettes was shown to be both effective and very cost-effective (29). However, in the development stages of the intervention (see appendix for details) both youth who had received cessation counseling and youth who had never tried any form of cessation counseling pointed out that face-to-face counseling would have a greater impact on their motivation to quit than online or automated text-generated counseling. With an increasing number of young people using tobacco and nicotine products, there will also be a greater demand for effective cessation counseling (1). Young people have more un-assisted quit attempts than adult smokers but have lower success rates (30). Therefore, youth who are unable to quit on their own should be offered the help they need and deserve.

## Data Availability

No datasets were generated or analysed during the current study. All relevant data from this study will be made available upon study completion.

## Statements and declarations

### Ethical considerations

The study is approved by the Danish National Scientific Ethics Committee (F-24044815).

### Consent to participate

Informed consent to participate will be obtained from all participants.

### Consent for publication

Not applicable.

### Data availability statement

A summary of the feasibility study and development of the U-Niko score is presented in the appendix. Data will be available upon reasonable request.

### Trial status

Recruitment of municipalities as clusters began March 25th, 2024, and ended April 23rd, 2024.

Recruitment of youth participants for cessation counseling began on January 1^st^, 2025, and ends on December 31^st^, 2025.

### Authors’ contributions

All authors have made substantial contributions to the development of the study protocol. SKBR and CP led the writing of the protocol, and MR, CBP, and NKKL provided critical insights into the development of the evaluation design and the writing of the study protocol, specifically the analysis plan (MR) and the methods section (CBP and NKKL). All authors have approved the submitted version of this paper and agree to be personally accountable for the author’s contributions and ensure that questions related to the accuracy or integrity of any part of the work, even ones in which the author was not personally involved, are appropriately investigated, resolved, and the resolution documented in the literature.

## Acknowledgments

The authors would like to express their sincere gratitude to Maj-Britt Bjerre Koch and Tine Vestergaard from Rygestopkonsulenterne ApS (The Smoking Cessation Consultants) for their invaluable expertise and collaboration in the development of the U-Niko intervention. Additionally, the authors wish to acknowledge the municipalities, counselors, schools, and students who participated in the development and testing of the U-Niko intervention.

